# Characterization of genomic regulation profiles in human mitral valve whole tissue to annotate genetic risk loci for mitral valve prolapse

**DOI:** 10.1101/2020.06.04.20122010

**Authors:** Sergiy Kyryachenko, Adrien Georges, Mengyao Yu, Takiy Barrandou, Patrick Bruneval, Tony Rubio, Judith Gronwald, Hassina Baraki, Ingo Kutschka, Russell A Norris, Niels Voigt, Nabila Bouatia-Naji

## Abstract

**Rationale:** Mitral valve prolapse (MVP) is a common valve disease that leads to mitral insufficiency, heart failure and sudden death. The identification of risk loci provided insight into its genetic architecture, although the causal variants and target genes need to be fully characterized.

**Objective:** To establish the chromatin accessibility profiles and gene regulation specificities of human mitral valve and identify functional variants and target genes at MVP loci.

**Methods and Results:** We mapped the open chromatin accessible regions in nuclei from 11 human mitral valves by an assay for transposase-accessible chromatin with high-throughput sequencing (ATAC-Seq). Compared to the heart tissue and cardiac fibroblasts, we found that mitral valve-specific ATAC-Seq peaks were enriched near genes involved in extracellular matrix organization, chondrocyte differentiation, and connective tissue development. The most enriched motif in mitral valve-specific open chromatin peaks was for the nuclear factor of activated T cells (NFATC) family of transcription factors, involved in valve endocardial and interstitial cells formation. We also found that MVP-associated variants (p < 10^−5^) observed in the current MVP GWAS were significantly enriched (p< 0.05) in mitral valve open chromatin peaks. Integration of the ATAC-Seq data with GWAS loci, extensive functional annotation, and gene reporter assay revealed plausible causal variants at two risk loci: rs6723013 at the *IGFBP5/TNS1* locus and rs2641440 at the *SMG6/SRR* locus. Circular chromosome conformation capture followed by high-throughput sequencing provided evidence for several target genes, including *SRR, HIC1*, and *DPH1* at the *SMG6/SRR* locus and further supported *TNS1* as the most likely target gene on Chr2.

**Conclusions:** Here we describe unprecedented genome-wide open chromatin profiles from human mitral valves that indicates specific gene regulation profiles, compared to the heart. We also report *in vitro* functional evidence for potential causal variants and target genes at MVP risk loci involving established and new biological mechanisms relevant to mitral valve disease.

## Introduction

Mitral valve presents specific cellular and tissue organization, compared to the heart and the vessels. The mature mitral valve is mainly composed of valvular interstitial cells (VICs), and is covered by a layer of valvular endothelial cells (VECs). These cells communicate via paracrine signaling, and altered or damaged signaling within the VECs can lead to pathological changes in underlying VICs.^1,2,3^ VICs are relatively quiescent non-contractile fibroblast-like cells that contribute to homeostasis of extracellular matrix (ECM).^4^ During valve development or under mechanical stress, VICs change phenotype and become activated myofibroblast-like cells that produce matrix metalloproteinases and inflammatory cytokines, which increase matrix production and remodel the ECM.^5,6,7^ Under excessive constant stress or TGF-β stimulation, this process leads to myxomatous degeneration of the valve causing mitral valve prolapse (MVP), a common valve disease. MVP affects nearly one in 40 adults and predisposes to mitral regurgitation, arrhythmias, and sudden cardiac death.^8,9,10^ Mitral regurgitation is the second most frequent indication for valve surgery, where valve reparation and replacement represent an important public health cost.^11^ The causes of MVP development and evolution are still poorly understood.

In a previous genome-wide association study (GWAS) for MVP, we identified six validated genetic risk loci, all located in intergenic or intronic noncoding regions.^12^ Associated variants are potentially located near regulatory elements that may be involved in the adjustment of target genes expression, specific to mitral valve. In the context of GWAS association signals, typically including large number of highly correlated genetic variants, the identification of potential causal variants and target genes requires the annotation of loci with epigenomic data generated from disease-relevant tissues.^13,14^ However, the most commonly used techniques, such as DNase-seq or histone-ChIP, require relatively high amount of cells, which excludes many tissues, including heart valve, from such analyses. On the other hand, expression quantitative trait loci (eQTL) in disease-relevant tissues are widely used to narrow down causal variants and potential target genes at GWAS loci,^15^ but such datasets are not available for the mitral valve.

Here we aimed to fill in the existing gap in functional annotation datasets for human mitral valve using the assay for transposase accessible chromatin using high-throughput sequencing (ATAC-Seq) approach, a genomic technique that provides a high-resolution map of open chromatin from samples with low cell content, as is the case of mitral valve tissue. Using this genomic annotation technique, we provide valuable information about active gene expression and main regulatory elements within human mitral valve cells. As a complementary approach, and in the absence of eQTL datasets for mitral valve, we performed circular chromatin conformation capture analyzed by high-throughput sequencing (4C-Seq) to provide evidence for physical proximity between MVP-associated variants and promoters as an indication of possible regulatory event between genetic variants and target genes.^16,17^ Finally, we provide two successful examples of the application of the genomic annotation, which we have generated, combined with chromatin architecture and gene reporter enhancer assay *in vitro* to propose candidate genetic variants for causality and their potential target genes at MVP GWAS loci.

## Materials and Methods

### Sample acquisition and patient consent

Mitral valve tissue was acquired at University Medical Center Göttingen, Georg-August University of Göttingen, Germany with written and informed consent through Institutional Review Board (IRB) protocol Nº 4/11/18. Freshly obtained mitral valve tissue from 11 patients who underwent mitral valve replacement surgery (Supplementary Table S1) was snap frozen in liquid nitrogen and stored at −80°C.

### Cell culture

Primary adult human dermal fibroblasts (HDF) (ATCC, Manassas, VA) and human cardiac fibroblasts (HCF) (Cell Applications, San Diego, CA) cells were purchased at passage 2 from LGC Standards, France and Tebu-bio, France respectively, and cultured in 5% CO2 in a 37°C incubator. HDF were maintained in fibroblast basal medium (ATCC, Manassas, VA) with fibroblast growth kit-low serum (ATCC, Manassas, VA), and HCF were maintained in HCF growth medium (Cell Applications, San Diego, CA) according to the manufactures’ instructions.

### Isolation of nuclei from frozen mitral valves for ATAC-Seq

Frozen mitral valve tissues were mechanically dissociated in liquid nitrogen using mortar and pestle. Grinded tissue was transferred into 15 ml Falcon tubes containing 5 ml of cold (Roche, Pleasanton, CA). The homogenate was filtered through 400 µm cell strainer homogenization buffer (HB): 10 mM Tris-HCl pH 7.6, 10 mM NaCl, 3 mM MgCl2, 0.1% NP40, 0.1% Tween-20, 0.01% Digitonin, 250mM Sucrose, and EDTA-free complete protease inhibitors (Roche, Pleasanton, CA). The homogenate was filtered through 400 μm cell strainer (pluriStrainer, Leipzig, Germany). Then, the isolation of nuclei was performed as described^18^ with modifications. Specifically, after Dounce homogenization, the tissue homogenate was pelleted at 500g for 10 minutes at 4°C. Tissue homogenate (700–800 μl) was transferred to a 14 ml round bottom tube (Greiner Bio-One, Frickenhausen, Germany) and mixed with an equal volume of 50% iodixanol in HB to obtain a final 25% iodixanol. 1200 μ l of 30% iodixanol in HB was layered underneath the 25% mixture, and then 1200 μl of 40% iodixanol in HB was layered below the 30% iodixanol. Nuclei were enriched at 3,000g for 20 minutes at 4°C in a swinging-bucket centrifuge. Nuclei that accumulated at the 30%/40% interface were collected into a 1.5 ml Lo-Bind Eppendorf tube, stained with Trypan Blue, and manually counted under microscope.

### Library preparation for ATAC-Seq

HDF and HCF cells at passage 5 were trypsinized, counted using a Nucleocounter NC-100 (Chemometec, Allerod, Denmark), and 50,000 cells were collected for each experiment. Isolated from mitral valves nuclei were transferred into a 1.5 ml Lo-Bind Eppendorf tube containing 1 ml ATAC-resuspension buffer with 0.1% Tween-20 and pelleted at 500 g for 10 minutes at 4°C.^18^ Depending on the number of nuclei (30,000–100,000) that were obtain, the transposition reaction was scaled up or down. The transposition reaction was performed as described in the Omni-ATAC protocol.^18^ Then, the transposition reaction was purified with MinElute PCR Purification Kit (QIAGEN, Hilden, Germany). Library amplification was performed as described previously.^19^ Amplified DNA was purified using Agencourt AMPure XP beads (Beckman Coulter, Brea, CA), according to manufacturer’s instructions.

### Sequencing of ATAC-Seq libraries and peak calling

ATAC-seq libraries were sequenced using 42 paired-end sequencing cycles on an Illumina NextSeq500 system at the high throughput sequencing core facility of Institute for Integrative Biology of the Cell (CNRS, France). Reads were demultiplexed using bcl2fastq2-2.18.12, and 31 to 95 million reads (fragments) were obtained per sample. Adapter sequences were trimmed using CutAdapt v1.15. For heart samples from ENCODE, raw reads from ATAC-Seq experiments on human heart left ventricle (ENCSR117PYB, ENCSR851EBF) and human right atrium auricular region (ENCSR062SVK) were downloaded from ENCODE webserver. Read length was adjusted to 42bp using Cutadapt. Further analyses were performed on the Galaxy webserver.^20^ Reads were mapped on GRCh38 (hg38) genome using Bowtie2 v2.3.4.3 with 150 default settings, except reads could be paired at up to 2kb distance. Aligned reads were filtered 151 using BAM filter v0.5.9, keeping only mapped, properly paired reads, and removing secondary 152 alignment and PCR duplicate reads as well as blacklisted regions.^21^ ATAC peaks were called 153 using MACS2 callpeak v2.1.1.20160309.6 with default settings. Binary read density files (bigwig) were created using bamCoverage v3.3.0.0.0, normalized on hg38 genome. BEDTool AnnotateBed v2.29.0 was used to compare peak files.

### Analysis of chromatin accessibility profiles

To perform sample correlation and principal component analysis, a common list of enriched regions was generated using bedtools multiple intersect (Galaxy Version 2.29.0) in “cluster” mode, and average read coverage on these regions was computed using deepTools multiBamsummary (Galaxy Version 3.3.2.0.0). deepTools plotCoverage and plotPCA functions were used to calculate Pearson correlation between samples and Principal Component Analysis, respectively. Global peak annotation was performed using ChIPSeeker v1.22.0.^22^ We used Diffbind (Galaxy Version 2.10.0) to detect differentially accessible regions between heart and mitral valve samples.^23^ clusterProfiler v3.14.0 was used to annotate genes at proximity of ATAC-Seq peaks and identify enriched gene ontology terms.^24^ Identified GOBP terms were clustered using REVIGO webserver (http://revigo.irb.hr/), with “medium” settings.^25^ We analyzed ATAC-seq peaks for enrichment in putative TF binding motifs using MEME-ChIP tool on MEME webserver (http://meme-suite.org/)^26^ set with HOCOMOCO Human v11 motif database. We used Integrated Genome Browser (IGB) to visualize read density profiles and peak positions in the context of human genome.^27^

### Prioritization of functional SNPs using annotation tools

Analysis of SNP enrichment among ATAC-Seq peaks was performed using GREGOR.^28^ The sentinel SNPs from loci associated with p-value < 10^−5^ were used as reference for MVP-associated SNPs. We included in the analysis SNPs in high LD (r^2^< 0.7) in European samples from 1000 Genomes. To use homogenous peak sets and mitigate the difference in signal-to-noise ratio between samples, 500bp windows centered on peak summits were used for this analysis. Next, to prioritize the most possible functional SNPs at each of the risk loci, we annotated each SNP with overlapping mitral valve ATAC-Seq peaks (narrowpeak from MCAS2 output + 100bp on each side), presence of overlapping H3K27ac histone mark from heart left ventricle, heart right atrium, ascending aorta or primary fibroblasts (peaks from ENCODE datasets: ENCFF052BXS, ENCFF172RFM, ENCFF208DZK, ENCFF222SST, ENCFF222WPT, ENCFF283CXG, ENCFF332RJQ, ENCFF385NNX, ENCFF401EEA, ENCFF491RWJ, ENCFF617BNF, ENCFF670ZUM, ENCFF686LUV, ENCFF824SPW, ENCFF946SRJ, ENCFF964UAX, and ENCFF984MJS) and RegulomeDB score.^29^

### Conditional analyses using GWAS data at *IGFBP5/TNS1* and *SMG6/SRR* loci

Using genotype data from our previous study^12^, we performed an updated association analysis on chromosome 2 and chromosome 17 using newly imputed SNPs obtained from the HRC^30^ as imputation reference panel through Michigan imputation server^31^. We used results from our GWAS meta-analysis as described previously.^12^ We used the GCTA-COJO function to perform conditional analyses to identify secondary or causal signals on loci.^32^ The COJO analysis used the MVP-France case control study as reference panel to calculate linkage disequilibrium (LD) between SNPs. To avoid collinearity issues, SNPs in high LD (r^2^>0.9) with the tested SNP were excluded before analyses.

### Circular chromatin conformation capture followed by high-throughput sequencing (4C-Seq)

The 4C template was prepared as previously described.^33^ In summary, 10 million of HDF were fixed in 2% formaldehyde, and then the cells were lysed to isolate nuclei. The chromatin was digested *in situ* with the four-base cutter MboI (New England Biolabs, Ipswich, MA) for 16 hours at 37°C followed by *in situ* ligation with T4 ligase (Thermo Fisher Scientific, Waltham, MA). Chromatin was decross-linked at 65°C overnight, followed by a second digestion with the four-base cutter NlaIII (New England Biolabs, Ipswich, MA). Digested fragments were diluted and self-ligated using T4 ligase to generate circular chromatin conformation capture (4C) library. The efficiency of each digestion and ligation step was validated on agarose gels. Viewpoints were selected on the basis of the MVP susceptibility loci found in the GWAS.^12^ To study the chromatin interactions of MVP-associated susceptibility loci with 4C, primers (Supplementary Table S2) were designed for each viewpoint as described previously.^33^ 4C libraries were sequenced using single-end 75bp reads on an Illumina NextSeq500 system at the high throughput sequencing core facility of Institute for Integrative Biology of the Cell (CNRS, France). The raw sequencing reads were demultiplexed on the basis of viewpoint-specific primer sequences. Reads were trimmed of sequences preceding MboI (GATC) site on 5’ end and following NlaIII site (CATG) on 3’end (if site was present in the read) using Cutadapt (Galaxy Version 1.16.6) on Galaxy webserver.^20^ Trimmed reads were mapped on GRCH37 (hg19, for r3Cseq package) and GRCH38 (hg38, for visualization) genomes using Bowtie2 v2.3.4.3 with default settings. Unmapped reads and secondary alignments were filtered out using Filter bam v0.5.9. Kept reads were processed using the R-package r3Cseq to detect significant interactions.^34^ All 4C-seq images were generated using default parameters of the pipeline.

### Vector construction, cell transfection, and dual-luciferase reporter gene assay

Details about primers and vector construction are presented in the supplementary appendix and in Supplementary Table S3. The constructed vector that contained the test SNP (90 ng for 96-well plate) and internal control plasmid pRL-TK (Promega, Madison, WI) (10 ng for 96-well plate) were co-transfected into HDF. The cells were plated at 4000 cells/well into a 96-well white plate 24 hours before transfection. The cells were transfected with a TransfeX transfection reagent (ATCC, Manassas, VA) according to the manufacturer’s instructions. Dual-luciferase reporter gene assays were performed after 48h post-transfection. Mithras LB 940 instrument (Berthold Technologies, Bad Wildbad, Germany) was used to measure the luciferase activity with the Dual-Luciferase Reporter Assay System (Promega, Madison, WI). All experiments were performed according to the instructions recommended by the manufacturer. The luciferase activity data were obtained from at least eight replicate wells, and the experiments were repeated 3 times. Two-tailed Student’s t-test was used to compare if the difference was significant and the significance threshold was set at P < 0.05.

## Results

### Mapping the chromatin accessibility profiles of human mitral valve tissue

We sought to get insight into the regulatory features of valve tissue by determining the chromatin accessibility profiles of surgically removed mitral valves using ATAC-Seq. We obtained between 55 and 95 million paired-end reads from the generated ATAC-Seq libraries of isolated nuclei from 11 valves. Peak calling ranged between 7,165 and 74,837 peaks, with five samples having at least 30,000 peaks (Figure 1A). We calculated Spearman correlation between samples to assess the variability in our experiment. We identified two main clusters: cluster A formed by the five samples where we obtained at least 30,000 peaks, and Cluster B formed by the six remaining samples, although these were relatively less correlated, indicating more inter-sample heterogeneity (Figure 1B). The visual examination of read density profiles showed that samples from Cluster A represented optimal quality ATAC-Seq data (Figure 1C). We therefore selected Cluster A samples (MV1671, MV1700, MV1772, MV1830, and MV1846) for further analyses.

**Figure 1.**
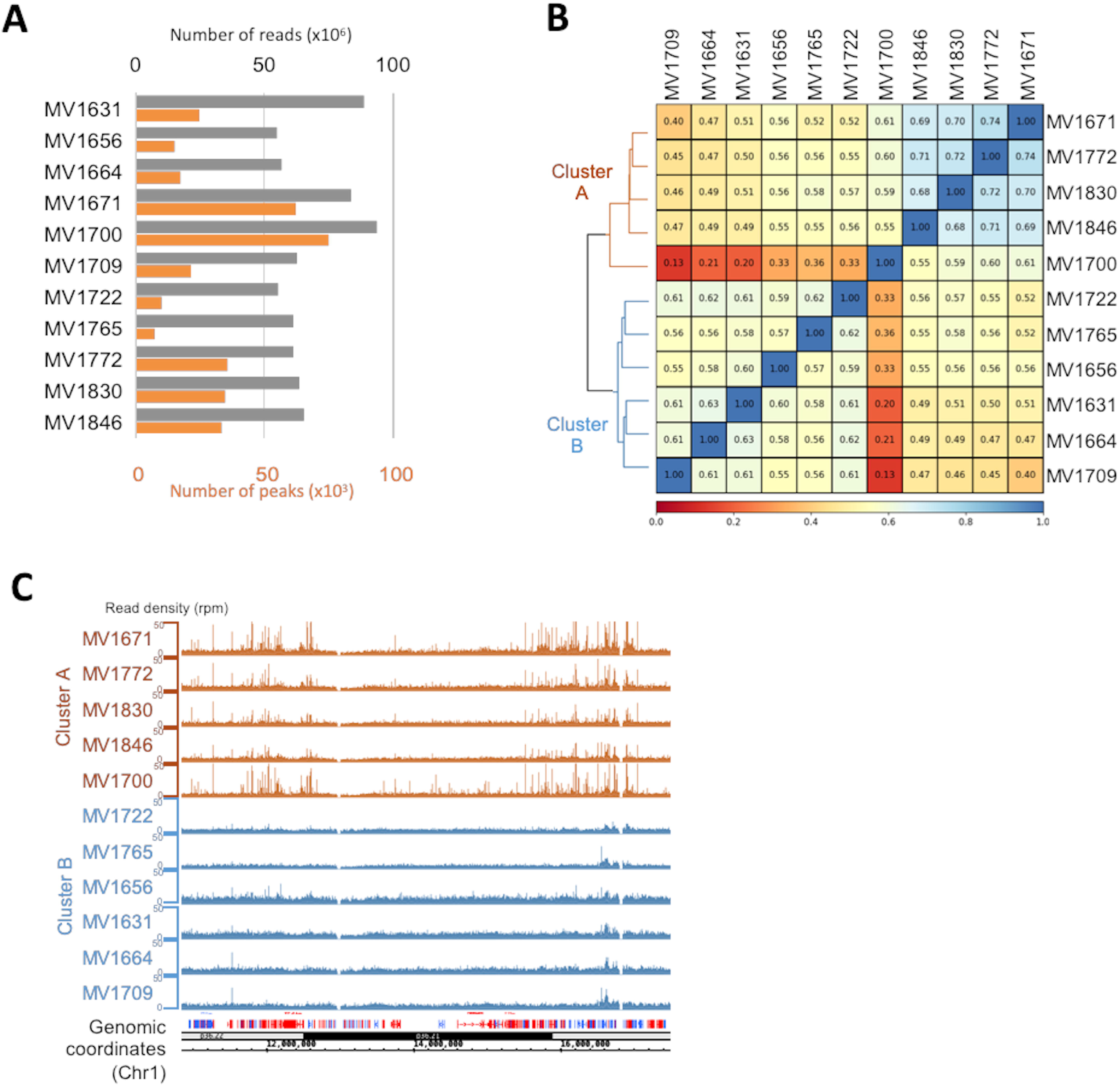
Quality controls of mitral valve ATAC-Seq experiments. **A**: Number of reads (grey) and number of peaks (orange) obtained for mitral valve ATAC-Seq libraries. **B**: Spearman correlation and hierarchical clustering of mitral valve ATAC-Seq datasets. **C**: Representative read density profile of mitral valve ATAC-Seq datasets on a gene rich portion of chr1 (11,000,000 to 17,500,000, hg38 coordinates).

### Comparison of chromatin accessibility profiles between mitral valve, heart tissues, and fibroblast cells

To determine whether mitral valve ATAC-Seq allows the identification of valve specific regulatory elements, we compared our data to primary cultures of fibroblasts and heart tissue. We determined chromatin accessibility in human dermal fibroblasts (HDF, two samples) and human cardiac fibroblasts (HCF, one sample), and reanalyzed through our pipeline ATAC-Seq experiments on heart left ventricle (Heart LV, two samples) and heart right atrium auricular region (Heart RAAR, one sample) from ENCODE. We obtained more than 130,000 peaks in all three fibroblasts samples, and 20,000 to 40,000 peaks from heart tissue (Figure 2A). We then annotated the genomic localization of ATAC-Seq peaks obtained in all samples. A large proportion of peaks observed in mitral valves were located in promoter regions (25 to 50%, according to samples, Figure 2B). Overall, 75 to 80% of peaks were located in the promoter or transcribed regions of genes, corresponding to usual localization of regulatory elements in the human genome identified by ATAC-Seq.^35^

**Figure 2.**
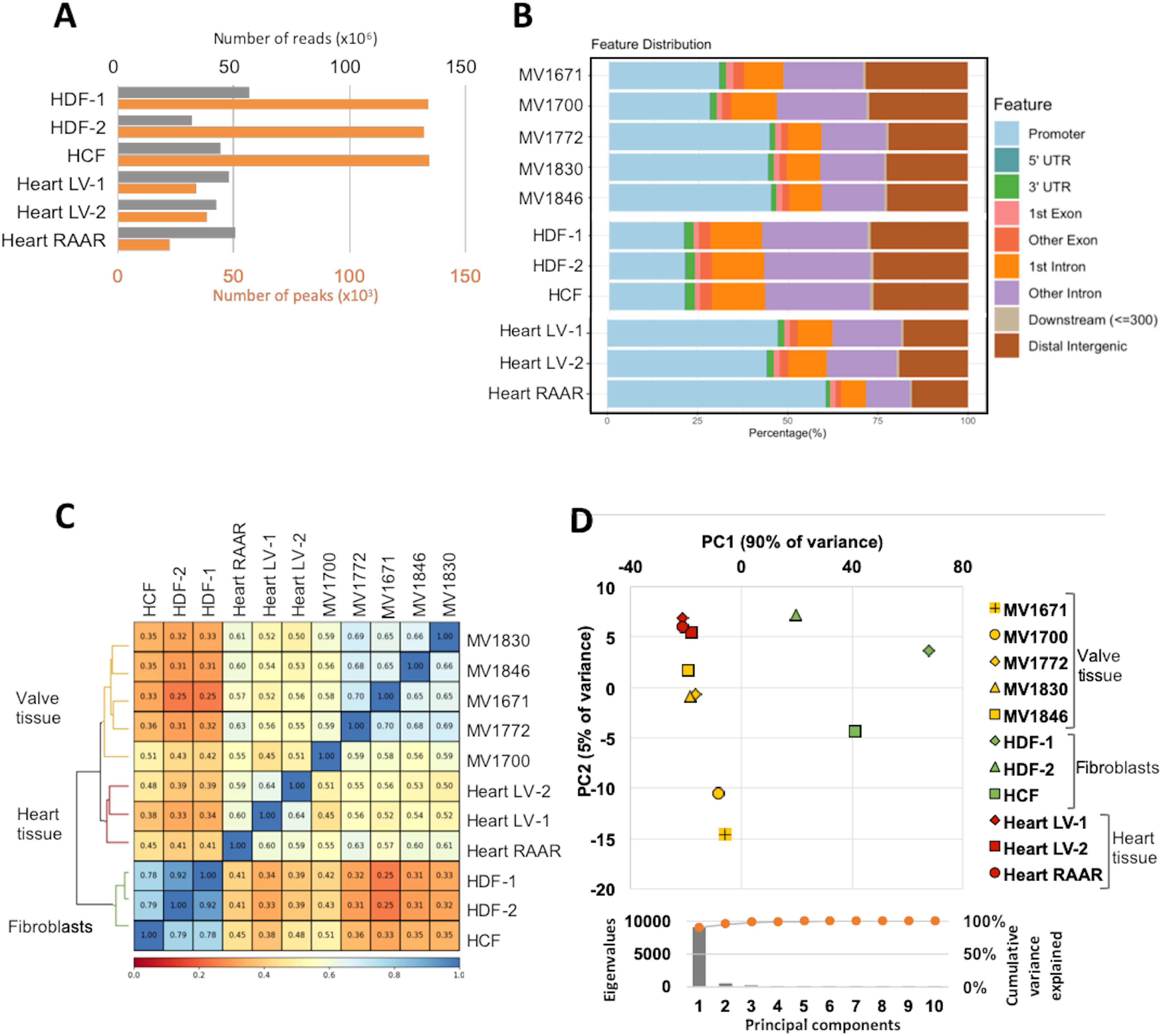
Comparison of mitral valve, heart, and fibroblast cells ATAC-Seq datasets. **A**: Number of reads (grey) and number of peaks (orange) obtained for HDF and HCF ATAC-Seq libraries as well as heart tissues: left ventricle (Heart LV) and right atrium auricular region (Heart RAAR) raw reads obtained from ENCODE database. **B**: Proportion of ATAC-Seq peaks located to different genomic regions. **C**: Spearman correlation and hierarchical clustering of ATAC-Seq datasets from 5 mitral valve samples, 3 fibroblasts samples, and 3 heart tissue samples from left ventricle (Heart LV) or right atrium auricular region (Heart RAAR). **D**: Principal Component Analysis of 5 mitral valve, 3 fibroblast, and 3 heart ATAC-Seq datasets. Upper panel shows the position of samples with respect to first 2 principal components. Lower panel indicates the eigenvalues and explained variance of the first 10 principal components.

We calculated Spearman correlation between all samples and identified three main clusters corresponding to mitral valve, heart samples, and primary cultured fibroblasts (Figure 2C). Heart samples from ENCODE clustered closer to mitral valve than to primary fibroblasts. Similarly, principal component analyses showed that tissue samples clustered together and separately from fibroblasts along the first principal component, which explains about 90% of the observed variance, whereas heart and mitral valve samples were separated by second principal component, accounting for about 5% of the total variance (Figure 2D).

### Chromatin accessibility profiles present regulatory features specific to mitral valve

To detect mitral valve-specific regulatory elements, we compared mitral valve samples to heart tissue samples. We identified 9758 peaks specifically enriched in the heart or mitral valve (FDR≤0.05, Figure 3A). To assess the global gene function controlled by these putative regulatory regions, we considered genes for which the transcription starting site (TSS) was located at 10kb or less distance from the enriched peaks (promoters or proximal regulatory elements), and we performed their functional annotation using Gene Ontology Biological Process (GOBP) terms. Genes at proximity of heart-specific ATAC-Seq peaks present strong enrichment for genes involved in heart contraction (p = 6×10^−19^), muscle cell differentiation (p = 3×10^−16^), and other pathways related to heart function (Figure 3B, Supplementary Table 1 and 2). Mitral valve-specific ATAC-Seq peaks were strongly enriched at proximity of genes involved in different biological pathways including ECM organization (p = 5×10^−15^), chondrocyte differentiation (p = 6×10^−11^), and terms associated to connective tissue development (Figure 3C, Supplementary Tables 3 and 4). We also note among top enriched terms those related to cell-cell adhesion and communication (e.g actin filament organization and the ephrin receptor signaling pathway) (Supplementary Tables 3 and 4).

**Figure 3.**
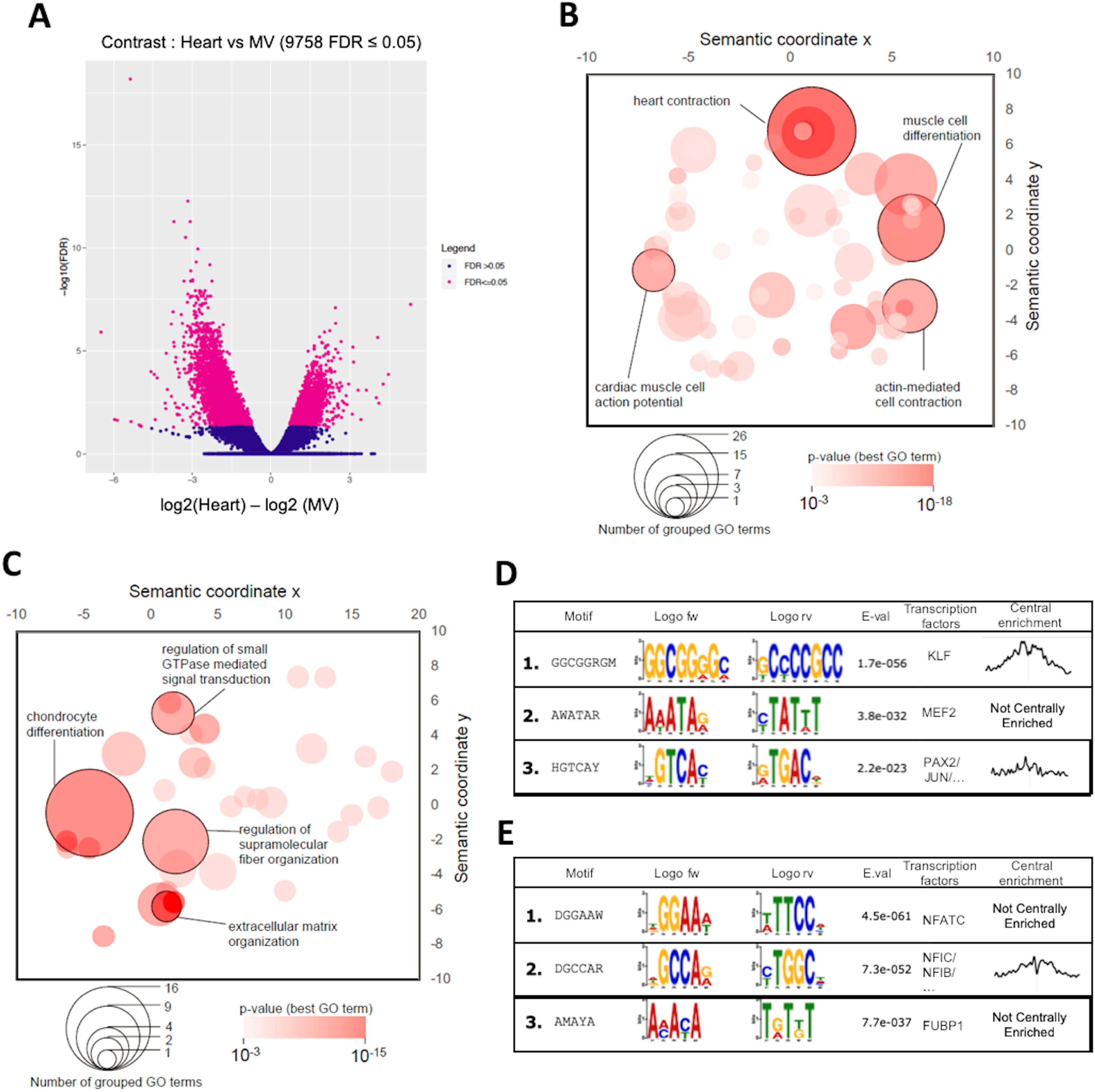
Analysis of ATAC-Seq identifies mitral valve-specific regulatory elements and pathways. **A**: Volcano plot representing negative logarithm of enrichment false discovery rate (FDR) on the Y axis and logarithm of enrichment in heart samples over mitral valve samples on the X axis. Each dot represents an ATAC-Seq peak in the mitral valve or heart sample. Pink dots represent significantly enriched regions in mitral valve or heart ATAC-Seq experiments (FDR≤0.05). **B-C**: Bubble graphs represent GOBP terms enriched among genes at proximity (distance to TSS≤10kb) of heart- (**B**) and mitral valve- (**C**) specific peaks. X and Y axes are in arbitrary semantic coordinates.^25^ Bubble size represents number of semantically similar terms aggregated under the same index GO term. Bubble color represents the p-value of GO term enrichment for the index GO term. **D-E**: Motif sequence and logo of most enriched motifs in heart- (**D**) and mitral valve- (**E**) specific peaks. Motifs were detected *de novo* using DREME algorithm. Top 3 motifs are represented. E-val indicates the erased expected value (E-value) calculated by DREME. “Transcription factors” indicate the top transcription factor or transcription factor family detected by TOMTOM algorithm as possible factors binding the detected motif. “Central enrichment” represents the enrichment of motifs with respect to ATAC-Seq peak summits.

Results of the enrichment analyses for transcription factor motifs also differ between heart and mitral valve. The most enriched motifs among heart-specific peaks were motifs for transcription factors involved in heart development and function^36,37,38^, mainly the Kruppel-like factor (KLF) and myocyte enhancer factor-2 (MEF2) families (Figure 3D). In the mitral valve-specific peaks, the top enriched motif corresponds to transcription factors of the nuclear factor of activated T cells (NFATC) family (Figure 3E).

### Mitral valve specific open chromatin peaks are enriched for MVP associated loci

Mitral valve ATAC-Seq data provide potential regulatory elements that may participate to the pathogenesis of MVP. To investigate further this idea, we tested the enrichment for associated SNPs from existing MVP GWAS results among different ATAC-Seq datasets.^12^ We used the Genomic Regulatory Elements and GWAS Overlap algorithm (GREGOR)^28^ that compares the number of trait-associated SNPs overlapping predefined genomic regions (here, ATAC-Seq peaks) with randomized SNP matched by regions (1Mb window) and frequency. Figure 4A shows that MVP-associated SNPs (p < 10^−5^) were significantly enriched (p < 0.05) in ATAC-Seq peaks from all mitral valve samples whereas significant enrichment was found for only one of the other samples (Heart LV-2). Maximal enrichment was seen for MV1830, for which ATAC-Seq peaks overlapped over 4 times more MVP-associated SNPs than expected (p = 6×10^−6^, Figure 4A). Comparatively, ATAC-Seq peaks from fibroblasts samples did not significantly overlap with MVP-associated SNPs, despite the fact that most peaks from mitral valves were also observed in fibroblast ATAC-Seq peaks.

**Figure 4.**
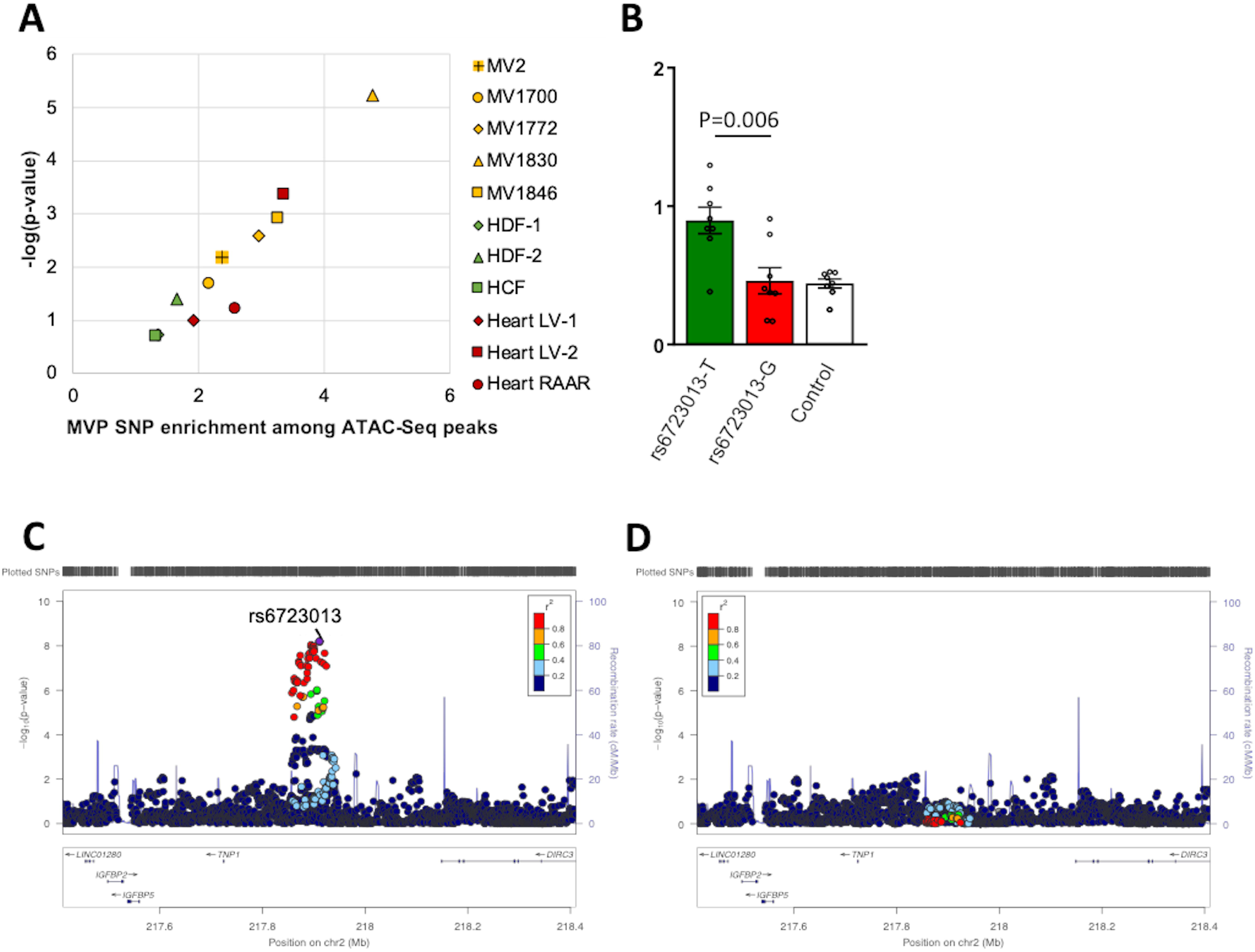
rs6723013 is a potential causal variant at the *IGFBP5/TNS1* MVP-associated locus. **A**: Representation of MVP SNPs fold-enrichment (X-axis) and enrichment p-value (log scale, Y-axis) among indicated ATAC-Seq samples. MVP SNPs overlap with ATAC-Seq peaks was compared to 500 pools of randomized matched SNPs to calculate the indicated enrichments. **B**: Luciferase reporter gene assay comparing the regulatory activity of constructs containing rs6723013-region (T and G alleles) and a control region. The p-value of a student’s t-test comparing luciferase values of rs6723013 alleles is indicated. **C-D**: Conditional analysis at *IGFBP5/TNS1* locus. LocusZoom plots represent MVP association before (**C**) and after (**D**) conditioning on rs6723013.

### Identification of potential causal variants at MVP-associated loci

Given the enrichment for regulatory elements near genes involved in myxomatous valve biology, we sought to use the mitral valve ATAC-Seq datasets to narrow down a list of potential causal SNPs at MVP-associated loci. We selected associated SNPs (p < 0.001) from the six loci reaching genome-wide significance in our previous study^12^ and filtered for colocalization with mitral valve ATAC-Seq peaks (Table 1). Between three and 20 SNPs were identified at each locus, representing a list of potential causal SNPs. We annotated these SNPs using RegulomeDB scores from 1 (most likely to affect TF binding and expression of a target gene) to 6 (least likely), and for the presence of H3K27ac enhancer/promoter marks in heart tissue (left ventricle or right atrium), ascending aorta, and dermal fibroblasts (Table 1).

**Table 1.**
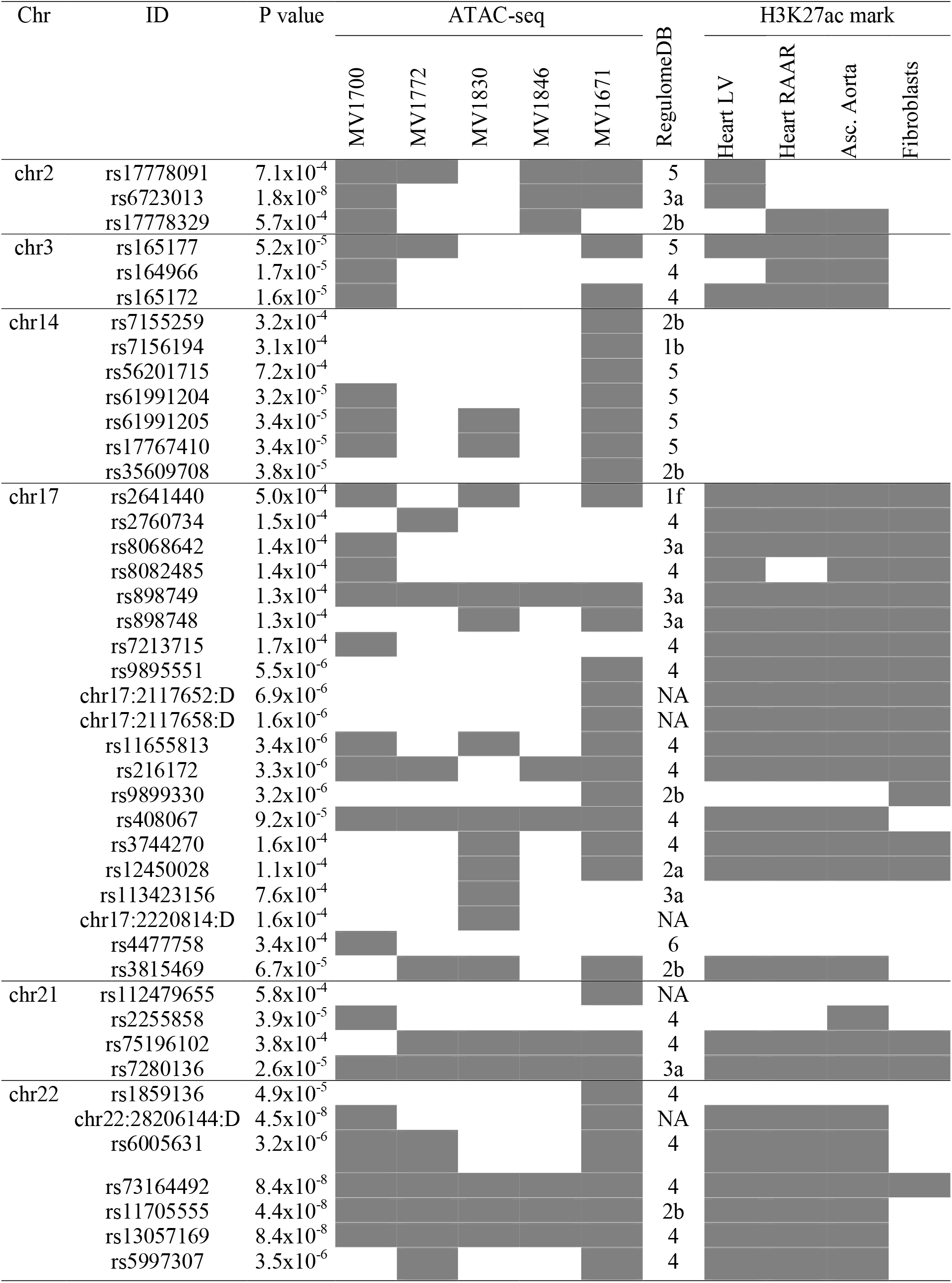
Potential causal SNPs at the MVP-associated loci. Overlaps with indicated epigenomic marks are indicated by grey cells. P values were reported for the association with MVP obtained in the discovery meta-analysis of GWAS^12^ NA: not-available.

To assess the validity of our approach, we sought to confirm the potential of selected SNPs as causal for MVP association. At the *IGFBP5/TNS1* locus on Chr2, where we had observed the strongest association signal in the discovery GWAS meta-analysis,^12^ only three SNPs overlapped ATAC-Seq peaks, and among them rs6723013 had the most significant association. Rs6723013 directly overlapped an H3K27ac histone mark in the heart left ventricle tissue, indicating it may be a part of an active enhancer region (Table 1). To further verify the regulatory effect of this SNP, we cloned 696bp DNA surrounding rs6723013, containing either rs6723013 reference (T) or alternative allele (G), upstream of *firefly luciferase* gene in a reporter vector. We then transfected these constructs in HDF, in which the region appeared to be active, and we measured the expression of luciferase. Interestingly, we found that T allele of rs6723013 was associated with significantly higher expression levels of firefly luciferase (reporter gene) than the alternative G allele of SNP rs6723013 (p = 0.006), whereas the construct containing G allele did not differ significantly from a control construct containing 751bp DNA from an inactive region (Figure 4B). These results show that rs6723013 genotype affects the regulatory properties of this enhancer and support it as a causal variant. In support of this, the conditional analysis on rs6723013, caused the association signal to totally disappear and indicated that this variant to totally drive the association at this locus (Supplementary Table S4, Figure 4C-D), (Figure 4D).

We applied the same strategy to the *SMG6/SRR* locus on Chr17, which is also known as a risk locus for other cardiovascular traits, such as aortic root size and coronary artery disease.^39,40,41^ Colocalization analysis showed that 21 SNPs directly overlap mitral valve ATAC-Seq peaks and 18 of these SNPs also overlap H3K27ac marks in at least one tissue, most of them in all the tested tissues. Among these SNPs, rs2641440 was ranked by RegulomeDB as the most likely regulatory SNP (RegulomeDB score = 1f), and it belongs to a strongly active region in all assayed tissues (Table 1). We also selected rs9899330 for *in vitro* tests, given its RegulomeDB high score 2b (Table 1), and low linkage disequilibrium with rs2641440 (r^2^ = 0.21, CEU 1000 genomes), suggesting it may belong to a secondary association signal. We then tested the effect of rs2641440 and rs9899330 genotypes on the regulatory activity of the surrounding region in HDF. The vector with the alternative C allele for rs2641440 had significantly higher luciferase activity compared with the construct containing the reference G allele (p = 0.0017) (Supplementary Figure S1A). Both constructs that contained alternative or reference allele of rs9899330 significantly enhanced the activity of luciferase compared with the control construct, but had no allelic effect on the regulatory activity of surrounding region (Supplementary Figure S1B). Interestingly, conditional analyses both on rs2641440 and rs9899330 at *SMG6/*SRR locus (Supplementary Table S4) caused a decrease of the association signal (Supplementary Figure S1C-F), supporting the existence of two independent signals at this locus.

### Identification of potential target genes at MVP-associated loci

To get further insight into the biology of MVP, we sought to determine potential target genes at each of the six confirmed MVP-associated loci^12^. First, we examined potential causal SNPs (Table 1) for eQTL association with genes in any human tissue using the Genotype-Tissue Expression (GTEx) portal (Table 2). Then, we performed circular chromatin conformation capture (4C) followed by high-throughput sequencing (4C-Seq) in human dermal fibroblasts to verify the potential target genes. We selected one to four viewpoints at each locus focusing on potentially functional SNPs according to the functional annotation, or at the closest genomic regions where we could design primers. Then, we used r3Cseq package to determine fragments that were significantly enriched in the 4C-Seq library (Supplementary Figure S2–5). Genes were considered as potential target genes if their promoter was present within 5kb of an enriched fragment or of a potentially functional SNP (Table 2). Neither eQTL nor 4C-seq analyses showed statistically significant association with target genes at the *IGFBP5/TNS1* MVP-associated locus (Supplementary Figure S2A, B, Table 2). At the Chr3 MVP-associated locus, we found interactions between the viewpoint and the promoter region of *LMCD1* gene (Supplementary Figure S2C) and no evidence for long range regulation for the nearby *CAV3*, which is highly expressed in the heart, and rare mutations may cause long QT syndrome.^42,43^ Our analyses also pointed out potential target genes at the Chr14, Chr21, and Chr22 MVP-associated loci, and further studies should explore their role in the MVP disease (Table 2, Supplementary Figure S3 and S5). On Chr17, variants at the *SMG6/SRR* locus had eQTL association with many potential target genes, including *SMG6, SRR, TSR1, SGSM2, HIC1*, and *DPH1* (Table 2). 4C-seq analyses from rs9899330 viewpoint showed significant interactions with promoters of *HIC1* and *DPH1*, which are located upstream of *SMG6*. (Figure 5, Supplementary Figure S4D). Interestingly, we detected numerous significant interactions between the selected viewpoints and the open chromatin regions within *SMG6/SRR* locus (Figure 5, Supplementary Figure S4). In particular, all viewpoints interacted with the same region containing strong enhancer signals close to a *SMG6* alternative promoter. These results suggest that active regions at this locus strongly interact together, and support the presence of multiple causal SNPs participating to the associations targeting multiple genes at this locus.

**Figure 5.**
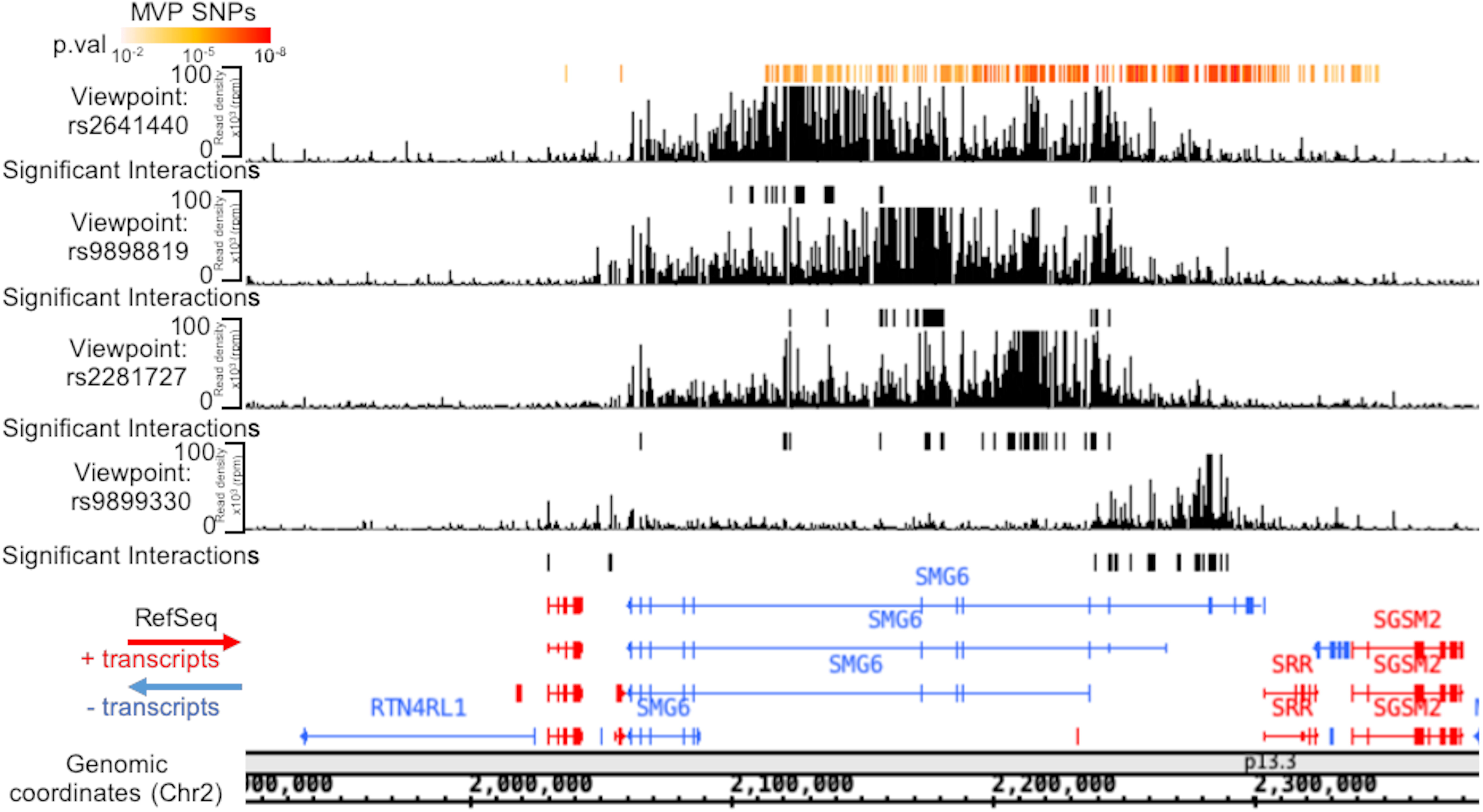
4C-Seq analysis of the *SMG6/SRR* MVP-associated locus. Interacting fragment density profiles from SNP rs2641440, rs9898819, rs2281727, and rs9899330 viewpoints using 4C-Seq. Fragments in significant interaction with the viewpoint are indicated by black bars under the interaction plot. Genomic coordinates, GRCh38.

**Table 2.**
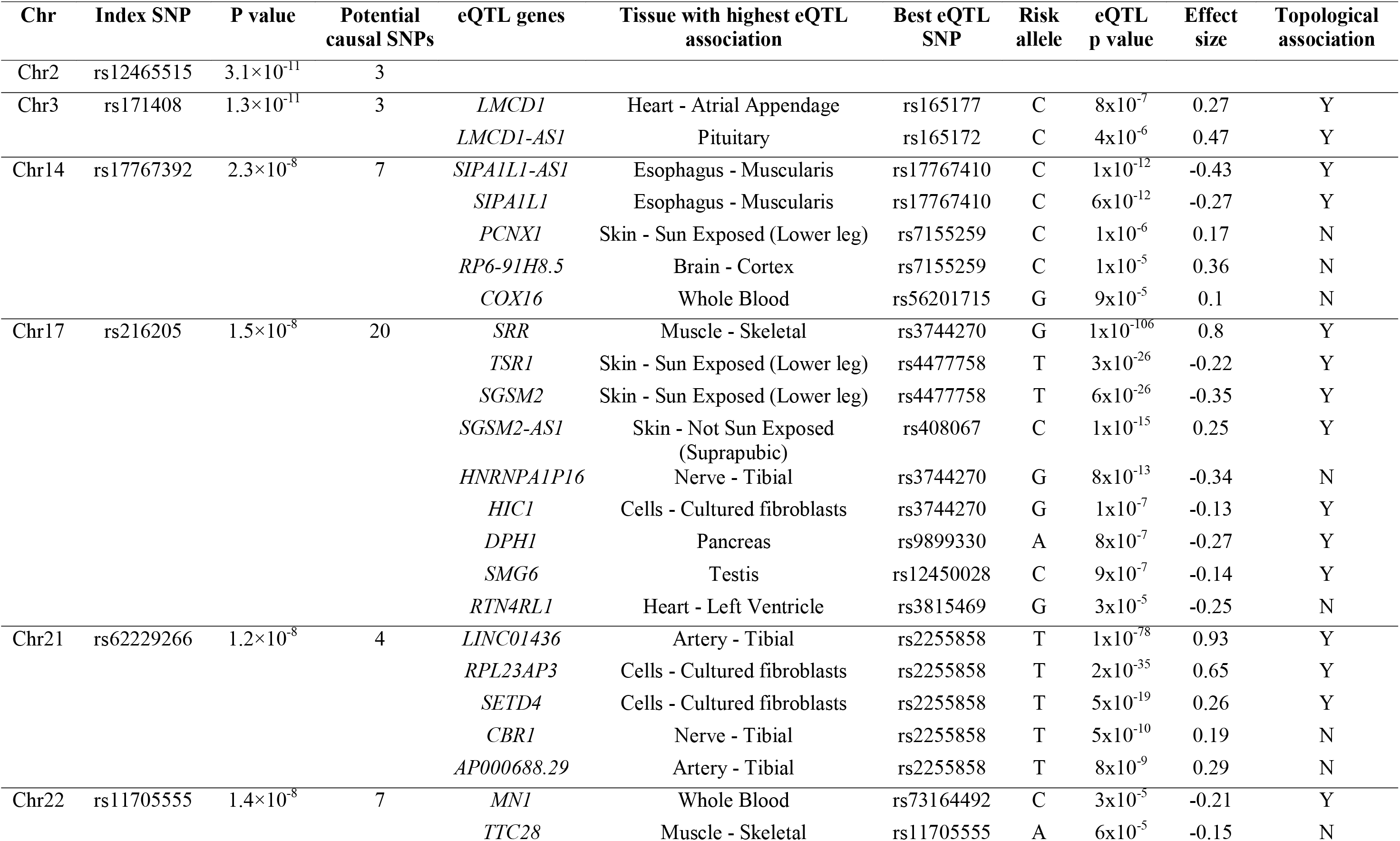
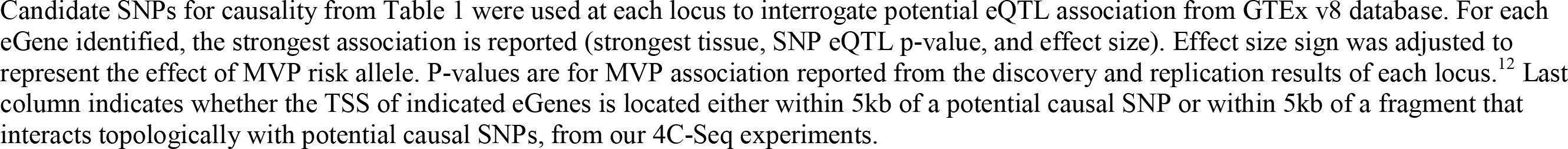
Potential target genes at MVP-associated loci.

## Discussion

In this study, we used epigenomic approaches to unravel the regulatory profile of human mitral valve tissue and leveraged this data to investigate functional variants and target genes at MVP-associated loci. We showed that mitral valves genome-wide open chromatin profiles are distinct from those of the cardiac tissue and fibroblasts, and are enriched near genes that are highly relevant and specific to the development of the valve and to the myxomatous process. Combining *in vitro* enhancer activity and chromatin tri-dimensional architecture, we were able to identify functional variants at the *IGFBP5/TNS1* and *SMG6/SRR* loci and narrow down the list of target genes at several MVP confirmed loci.

Our data support that open chromatin mapping of diseased mitral valves provides a useful resource on cell differentiation and regulation of the myxomatous process. We found that ATAC-Seq peaks were highly enriched for genes relevant to valve biology and disease, such as ECM and connective tissue development and cytoskeleton organization, including actin filament and cell adhesion. Of particular interest are the open chromatin peaks near genes related to the ephrin receptor signaling pathway, an important family of transmembrane tyrosine kinase receptors that ensure short cell-cell communication. This pathway is involved in the regulation of epithelial-to-mesenchymal transformation during heart valve morphogenesis, and *Efna1* knockout mice exhibit the thickening of both aortic and mitral valves.^44^ Another relevant process is chondrogenic differentiation, which is part of the valve myxomatous degeneration process of VICs.^45^ We found the NFATC family of transcription factors to be the most enriched motif in mitral valve-specific open chromatin regions. NFATC1, in particular, is known to be a key factor in heart valve and VICs development and its regulation may play a role in myxomatous degeneration.^46,47,48^ NFATC1 is also expressed in adult human pulmonary VECs, and VEGF-induced NFATC1 activation can participate in the valve maintenance by increasing proliferation of VECs.^49^ A study reported microarray gene expression to be increased for NFATC2 and NFATC4 in myxomatous mitral valves compared to normal valves.^50^ Our results encourage future studies to understand the specific role of NFATCs in the establishment of MVP.

Our study shows that MVP-associated SNPs are significantly enriched among identified mitral valve open chromatin peaks, but not among regions accessible in dermal or cardiac fibroblasts. Despite the fibroblast-like phenotype of VICs and VECs, our results are in favor of open chromatin and active regulation in mitral valve tissue to significantly differ from the one observed in cultured fibroblasts derived from the skin or the heart.

Our approach to fine-map GWAS loci using chromatin accessibility data derived from mitral valve tissue, combined with the visualization of long-range genomic interaction using 4C-Seq method, allowed us to identify potential causal variants and to narrow down the list of target genes at risk loci, especially on chromosomes 2 and 17. At the *IGFBP5/TNS1* MVP-associated locus we identified rs6723013 as a functional enhancer *in vitro* and demonstrated regulatory allelic effect. Presence of the minor T allele (T = 0.24, frequency in TOPMed panel) can modulate the expression of potential target genes by altering transcription factor binding onto DNA. Several TFs, such as OVOL2 and NFE2L1, are predicted to bind more to the major G allele. OVOL2 suppresses epithelial-to-mesenchymal transition and thus maintains epithelial lineages, and NFE2L1 regulates expression of target genes in response to various stresses. Further studies are needed to explore their involvement in gene regulation by this enhancer region. Potential target genes at this locus were *IGFBP5, IGFBP2*, and *TNP1* that are located upstream from the signal, and *DIRC1* and *TNS1* located downstream from the signal. According to the chromatin organization observed from our 4C-Seq data, *IGFBP2, IGFBP5*, and *TNS1* were all part of the same TAD, which argues for high probability of contact between these genes promoters and the enhancer containing the functional variant. *IGFBP2* and *IGFBP5* are two members of the insulin growth factor binding protein family. Igfbp5 is expressed in the developing mouse heart,^51^ and differentially expressed in VECs during postnatal ECM remodeling and leaflet morphogenesis in P7 and P30 murine valves.^52^ However, we have previously shown the absence of atrioventricular regurgitation properties in zebrafish knockdown experiments of *igfbp2* and *igfbp5* fish orthologs.^12^ In the same study, significant atrioventricular regurgitation was found in zebrafish knockdown, and *Tns1*^−/−^ mice exhibited myxomatous phenotype of mitral valves.^12^ In light of these elements, we conclude that *TNS1* is the most plausible causal gene at the *IGFBP5/TNS1* locus through a potential long-range gene regulation mechanism, similar to previously described genomic organizations.^53,54^

The *SMG6/SRR* locus is an intriguing and genetically complex locus reported to be associated with several cardiovascular traits, including coronary artery disease.^40,41^ Our updated association analysis, using a more dense imputed set of variants, provides several hundreds of variants in high LD to significantly associate with MVP at this locus. Filtering for the overlap with ATAC-Seq peaks in mitral valves and enhancer marks, prior to reporter assay experiments, provided evidence for enhancer capacity *in vitro* for rs2641440, with the G allelic variant (G = 0.64, TOPMed) at risk for MVP. Nonetheless, the conditional analysis supported the presence of several secondary signals at this locus, and we cannot exclude additional functional effects from the untested SNPs. The *SMG6/SRR* locus is located in a gene-rich region that contains overlapping promoters. Our 4C-Seq showed that all fours viewpoints that we tested as potential enhancers were located within open chromatin in mitral valves, suggesting synergic regulation of several nearby genes.^55^ We have previously excluded *SMG6* and *SGSM2* candidacy for causality at this locus as neither *smg6* nor *sgsm2* knockdown in zebrafish led to an abnormal cardiac phenotype.^12^ Our chromatin interaction results now highlight new potential target genes, including HIC ZBTB transcriptional repressor 1 (*HIC1*), diphthamide biosynthesis 1 (*DPH1*), and serine racemase (*SRR)* genes, all expressed in heart tissues.^56,57,58^ HIC1 is a partner of TCF-4, and this complex attenuates Wnt/β-catenin signaling, an important mechanism for heart valve development and myxomatous degeneration.^59,45^ On the other hand, patients with mutations in *DPH1* present heart abnormalities, including ventricular septal defect and aortic stenosis.^60^ Finally, several associated variants are located at the promoter of *SRR*, which catalyzes the synthesis of D-serine from L-serine, and among its related pathways is calcium signaling.^61^

Our work presents several limitations. First, it is the absence of non-pathogenic valves as controls, which limited direct comparison of the regulatory profiles between healthy and myxomatous valves. Second, open chromatin profiles obtained from ATAC-seq methods provide indirect assessment of transcriptional activity. Direct assessment of transcription using RNA-Seq would be useful to determine the specific regulatory features of mitral valves, although their poor cellular content represents a technical challenge. Third, enhancer activity tested *in vitro* in dermal fibroblasts may differ from *in vivo* native chromatin activity in VICs and VECs. Finally, our results are conditioned by the limited resolution of the 4C-Seq experiments, which do not allow refinement at gene-rich loci as the *SMG6/SGSM2*.

In conclusion, we provide for the first time mitral valve genome-wide chromatin accessibility profiles and demonstrate that this valuable data source to substantially contribute to deciphering MVP etiology. We show that MVP-associated variants are significantly and specifically enriched in open chromatin of myxomatous valves. Using *in vitro* functional validation, in combination with long range chromatin interaction experiments, we provide evidence for causal SNPs at two MVP-associated loci and several target genes of high relevance to valve development and degenerative process.

## Data Availability

ATACSeq data will be available through the repository Gene Expression Omnibus.

## ACKNOWLEDGMENTS

The authors thank Ines Müller and Stefanie Kestel for excellent technical assistance.

---

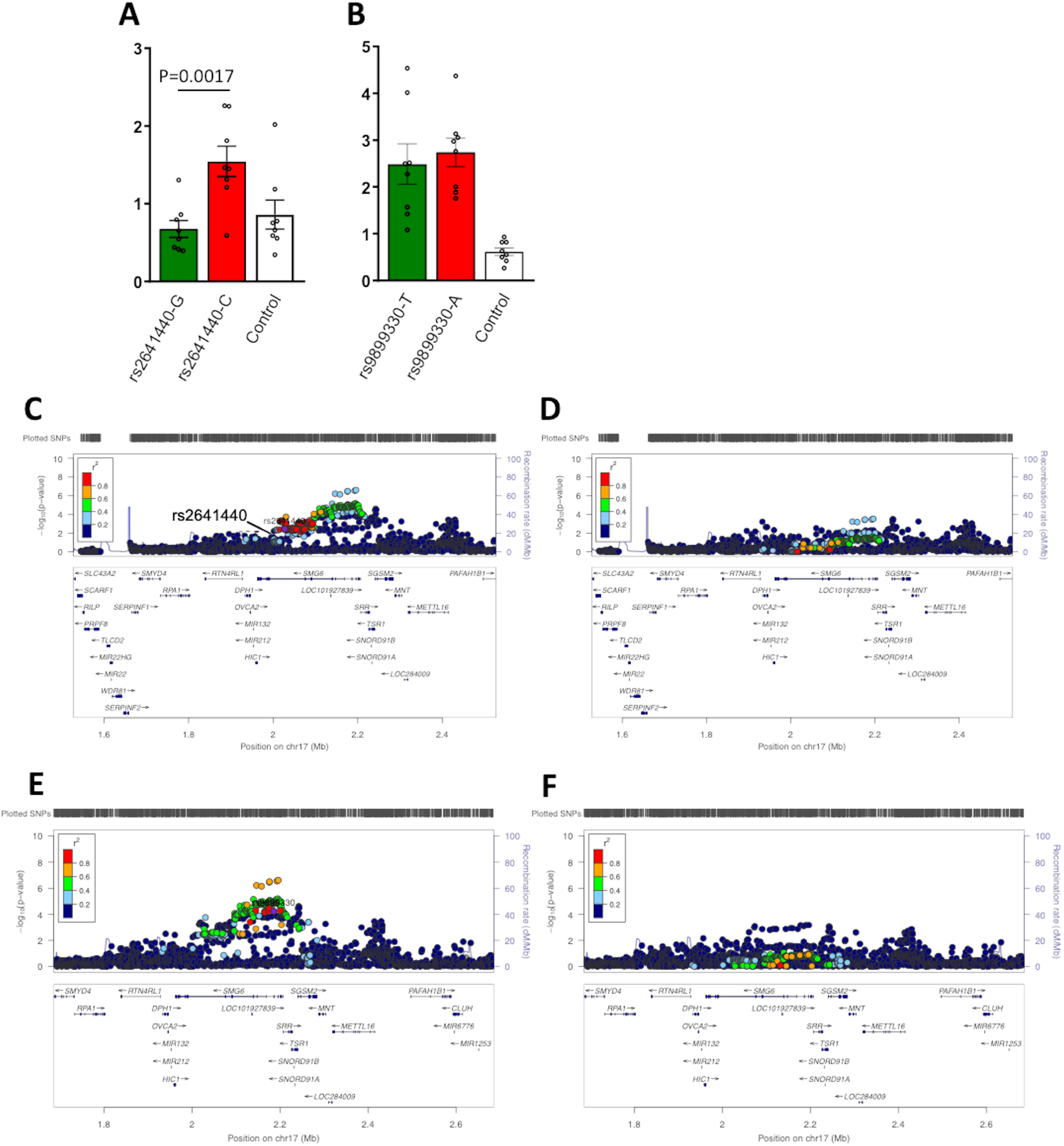
Fig S1

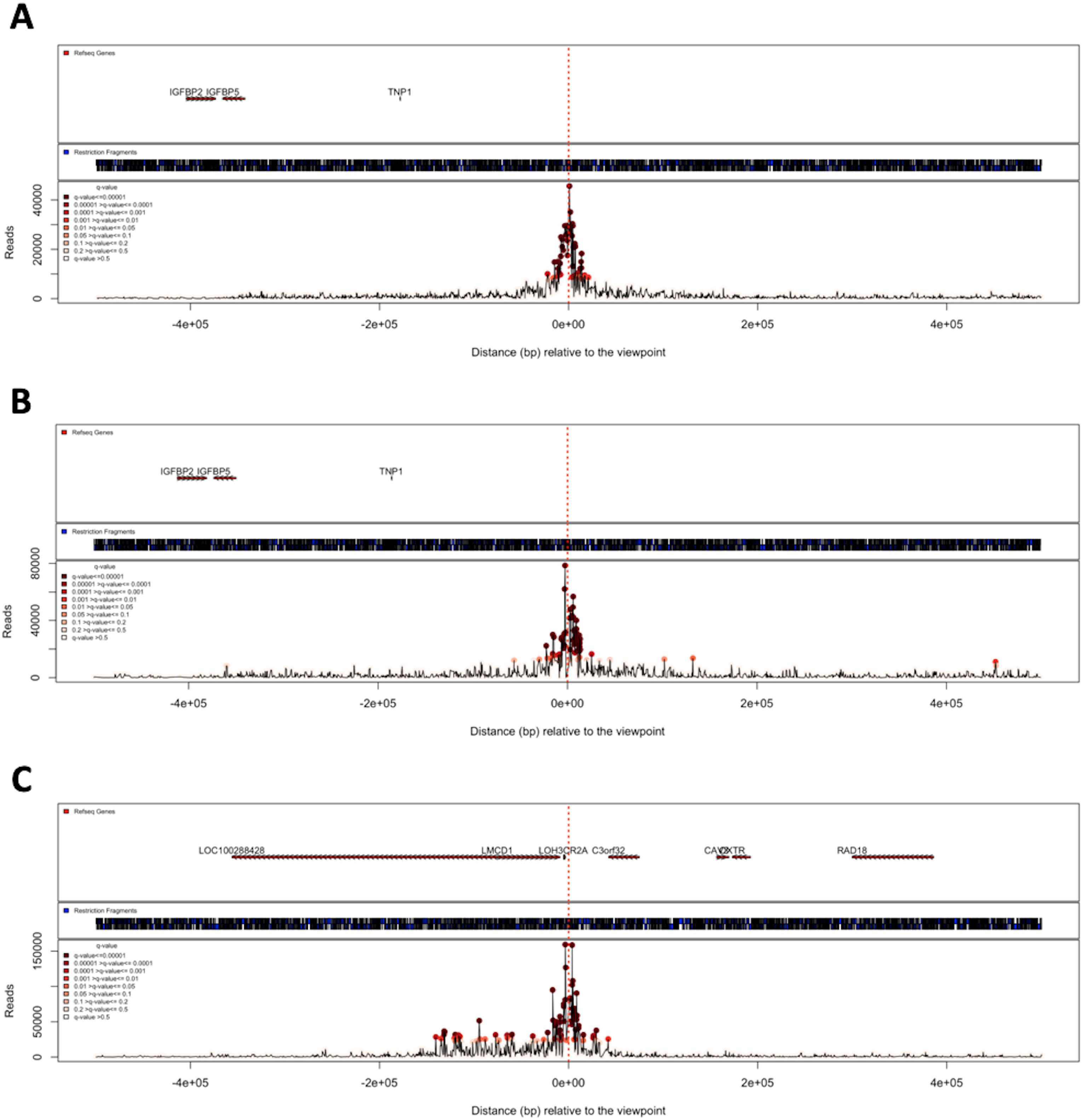
Fig S2

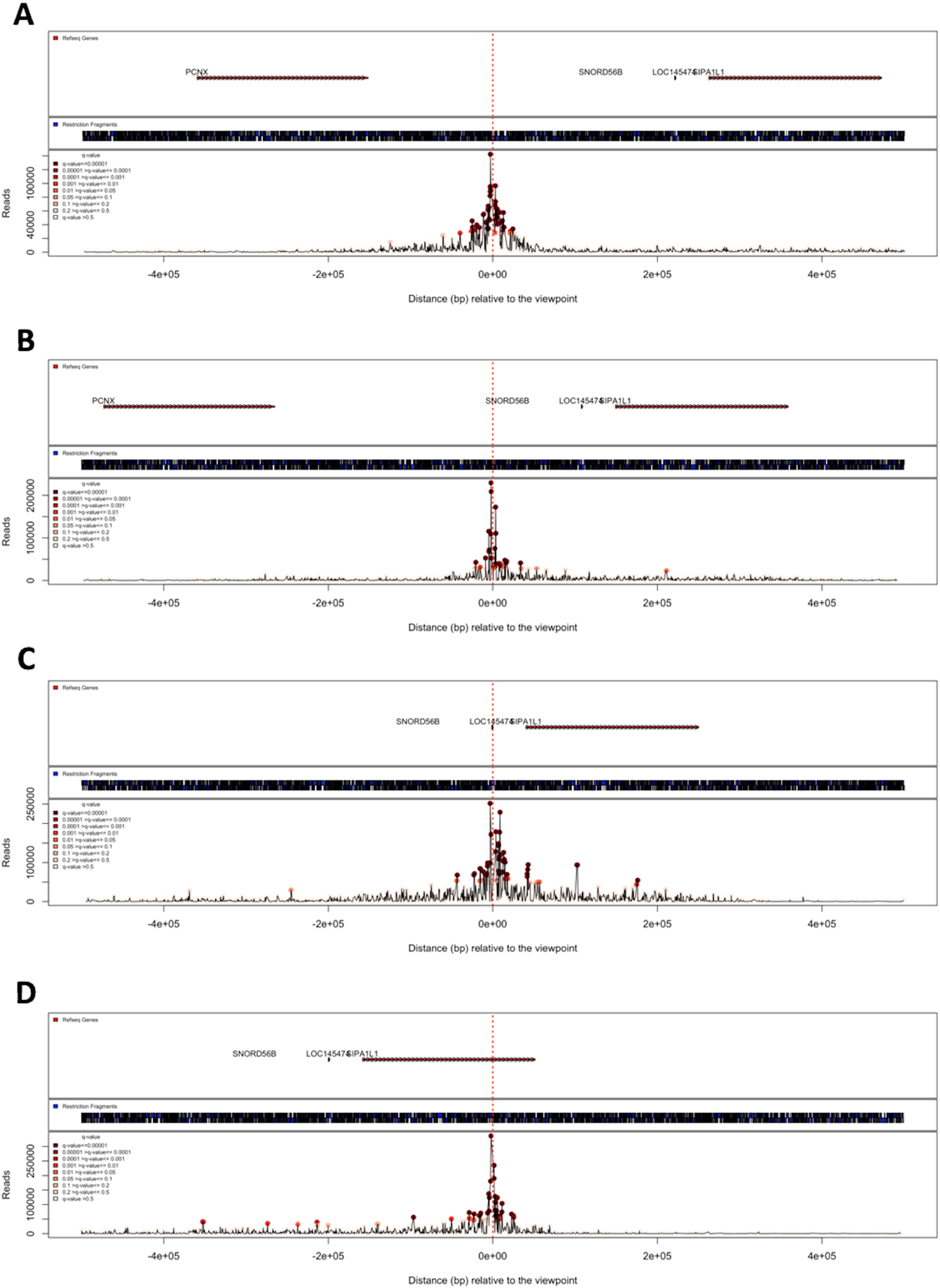
Fig S3

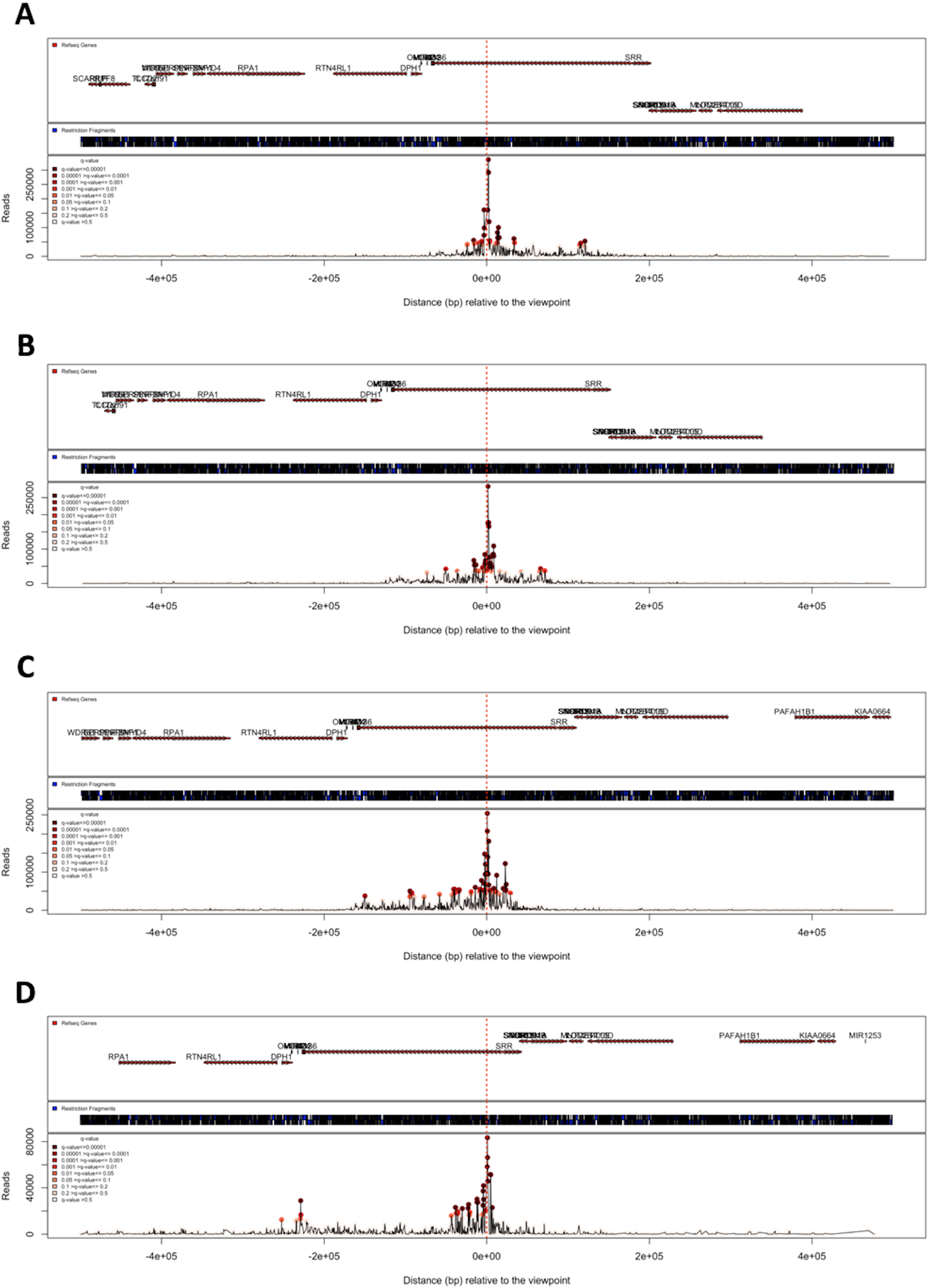
Fig S4

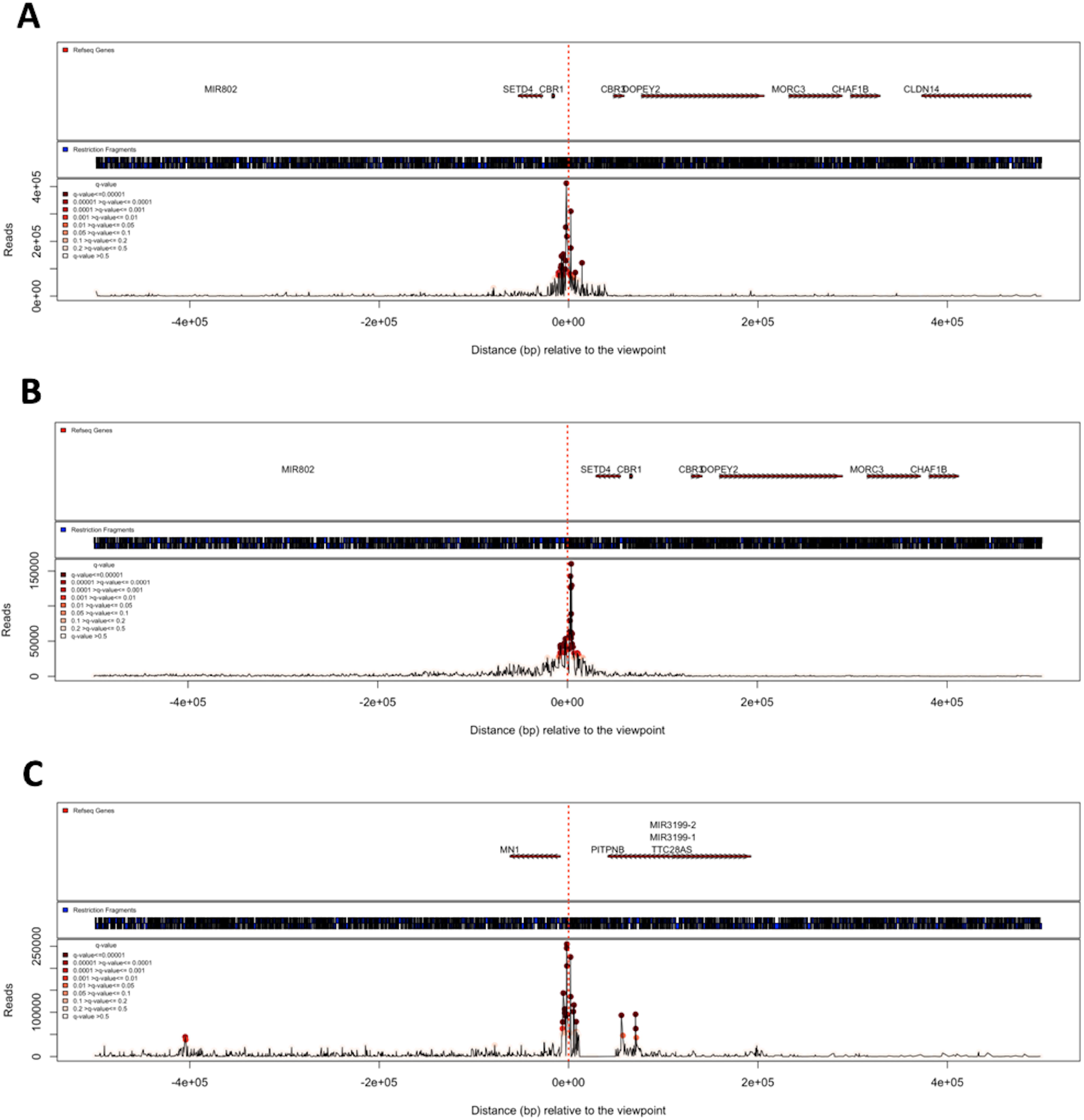
Fig S5

